# Asthma exacerbations and eosinophilia in the UK Biobank: a Genome-Wide Association Study

**DOI:** 10.1101/2023.04.12.23288479

**Authors:** Ahmed Edris, Kirsten Voorhies, Sharon M. Lutz, Carlos Iribarren, Ian Hall, Ann Chen Wu, Martin Tobin, Katherine Fawcett, Lies Lahousse

## Abstract

Asthma exacerbations reflect disease severity, affect morbidity and mortality, and may lead to declining lung function. Inflammatory endotypes (e.g.:T2-high (eosinophilic)) may play a key role in asthma exacerbations. We aimed to assess whether genetic susceptibility underlies asthma exacerbation risk and additionally tested for an interaction between genetic variants and eosinophilia on exacerbation risk.

UK Biobank data were used to perform a GWAS study of individuals with asthma and at least one exacerbation compared to individuals with asthma and no history of exacerbations. Individuals with asthma were identified using self-reported data, hospitalization data and General Practitioners (GP) records. Exacerbations were identified as either asthma–related hospitalization, GP record of asthma exacerbation, or an oral corticosteroid (OCS) burst prescription. A logistic regression model adjusted for age, sex, smoking status, and genetic ancestry via principal components was used to assess the association between genetic variants and asthma exacerbations. We sought replication for suggestive associations (P<5x10^-6^) in the GERA cohort.

In the UK Biobank, we identified 11,604 cases, and 37,890 controls. While no variants reached genome wide significance (P<5x10^-8^) in the primary analysis, 116 signals were suggestively significant (P<5x10^-6^). In GERA, two SNPs (rs34643691 and rs149721630) were nominally significant and showed the same direction of effect.

Two novel genetic loci-(NTRK3 and ABCA13)-that are reproducibly associated with asthma exacerbation in participants with asthma were identified. Confirmation of these findings in different asthma (or ancestry) sub-populations and functional investigation will be required to understand their mechanisms of action and potentially inform therapeutic development.

## Introduction

Asthma is a heterogeneous chronic respiratory condition, estimated to affect more than 300 million people worldwide.[1] The disease features a network of complex inflammatory phenotypes. Additionally, asthma has a well-established genetic component, associated with variability in its clinical presentation and asthma severity.[2] Several clinical phenotypes have been defined based on the onset of asthma, control of symptoms, and co-morbidity of allergy involved in the underlying pathophysiology.[3] In early attempts to disentangle the molecular pathophysiology of asthma, the disease was broadly divided into two major endotypes: type 2 asthma and non-type 2 asthma.[4]

The type 2 inflammatory endotype represents an important clinical challenge, as it is characterized by airway eosinophilia, difficulty to achieve asthma control, and a higher frequency of exacerbations.[4] [5] Moreover, a crucial subgroup of individuals with asthma with this endotype are uncontrolled despite treatment with inhaled corticosteroids. This group represent a large proportion of individuals with severe uncontrolled asthma and frequent exacerbations.[4] Type 2 inflammation is driven by increased activity of T-helper cells type 2 (Th2), activated by dendritic cells.[4] These secrete interleukin (IL)-5, IL-4 and IL-13 and other type 2 cytokines, which activate type 2 immunity pathways.[6, 7] Therefore, type 2 asthma is characterized by airway and systemic eosinophilia.[4] Type 2 development is thought to be driven by genetics, as well as epigenetic and environmental factors.[4, 8] Interestingly, Tantisira et al. show that genetic factors determining susceptibility to asthma differ from those determining its severity.[9] Determining the most important genetic variants affecting asthma severity may be fundamental to understand drivers of disease activity in asthma[10], and subsequently improve available treatment approaches, either by early prediction of severity, or finding new druggable targets for severe, uncontrolled asthma. Although several studies have attempted to disentangle the underlying genetic variants behind asthma, power and sample sizes required have necessitated the combination of samples including different potential asthma phenotypes, leading to increased heterogeneity.[2] Importantly, patients with type 2 asthma often find it difficult to achieve asthma control, and have frequent exacerbations.[5] Moreover, few studies have examined the genetic variants contributing to the severity of asthma within patients with asthma.

Asthma exacerbations are an important cause of asthma morbidity, mortality and healthcare costs.[11-15] Importantly, exacerbations are also useful in evaluating treatment response, and are a marker of asthma control.[16] Moreover, exacerbations may increase the rate of lung function decline, thus representing a clinically important long term outcome.[17, 18] Asthma exacerbations are known to be affected in part by genetics and epigenetics[2, 19], and are associated with the active inflammatory endotype.[5] Studies investigating asthma exacerbations focused mainly on hospitalizations and emergency department (ED) visits, and revealed several genes including *IL13, IL4RA, CHI3L1, ORMDL3, CDHR3, CTNNA3, SEMA3D, EXTL2* and *PANK1*.[9, 20] However, most studies investigating hospitalizations in asthma focused on childhood asthma, and only included a small number of events or cases.[9, 20, 21] An analysis in the UK Biobank focused on asthma hospitalizations (but not data from primary care records) and implicated genes in the HLA region.[22] In this study, we aimed to evaluate the genetic factors affecting asthma exacerbations in a large, genome-wide study. Additionally, we aimed to investigate whether eosinophilia modifies the effect of variants on exacerbations.

## Methods

### Study population

This study used a case-control design in 2 stages. The data source for the primary analysis was the UK Biobank (http://www.ukbiobank.ac.uk) and analysis was performed under approved application 648. The UK Biobank is a population-based study of half a million volunteer participants between the ages of 40 and 69 years, recruited from Great Britain in 2006-2010. In total, 321,057 individuals with genetic data were eligible for inclusion in this analysis. Data source for the replication was the GERA (Genetic Epidemiology Research in Aging) cohort. GERA is a multi-ethnic cohort of over 110,000 subjects from the Kaiser Permanente Medical Care Plan, Northern California Region (KPNC), Research Program on Genes, Environment, and Health who provided a saliva sample.[23] All races and ethnicities were self-reported, and 80% of subjects were non-Hispanic white. The GERA study was approved by the institutional review boards at KPNC and Brigham and Women’s Hospital (2002P000331).

### Definition of asthma

In the UK Biobank, asthma was defined as either: self-reported asthma in the touchscreen questionnaire (data field: 6152), an asthma code in general practitioner records (Read v2 and Read v3 codes, full list of codes is provided as supplementary tables S1 and S2, the choice of code was based on Mukherjee et al.[24]), or any hospitalization event with an asthma ICD10 code as cause for admission (ICD10 codes: J45, J45.0, J45.1, J45.8, J45.9, J46, J46.0). In GERA, over 16,000 asthmatics have genotype data and longitudinal electronic medical records, including detailed diagnosis and medication records. Prescription data was available from outpatient and emergency department visits. Adult asthma cases were defined as patients at least 21 years of age with one or more of the following: physician-diagnosed asthma, self-reported asthma, or a report of an asthma exacerbation (i.e., emergency department visit or hospitalization due to asthma).

### Exclusion criteria

In the UK Biobank, individuals were excluded from this study if they had incomplete genotyping data that did not pass quality control (as described in Shrine et al.[25] and Guyatt et al.[26]), non-European ancestry based on K-means clustering after principal component analysis (as described in Shrine et al.[25] and Guyatt et al. [26]), evidence of COPD (defined as either: self-reported COPD in the touchscreen questionnaire; a COPD hospitalization code (ICD10 codes: J44, J44.0, J44.1, J44.8, J44.9) or an FEV_1_/FVC ratio <0.7); evidence of chronic bronchitis and/or emphysema (defined as either: self-reported chronic bronchitis or emphysema in the touchscreen questionnaire or an emphysema hospitalization code: J43.2, J43.8, J43.9). Related individuals up to second degree relatives were excluded based on kinship coefficients using KING software.[27] In GERA, subjects with chronic obstructive pulmonary disease (COPD), pulmonary embolism, primary pulmonary hypertension, cystic fibrosis, emphysema, and chronic bronchitis bronchiectasis were excluded.

### Definition of asthma exacerbators

Individuals were classified as exacerbators using three different data sources: hospitalization data, primary care general practitioner records data, and primary care prescription data. Hospitalizations were considered the result of an asthma exacerbation if: (i) asthma was listed as a primary cause for hospitalization, (ii) asthma was listed as a secondary cause for hospitalization with the primary cause being a respiratory infection or condition associated with asthma (codes: J10.0, J10.1, J11.1, J11.8, J20.9, J67.9, J96.0, J96.00, J96.01, J96.09, J96.1, J96.11, J96.19, J96.9, J96.90, J96.91, J96.99, R06.1, R06.2, R06.4, R06.5), (iii) asthma was listed a secondary cause and the primary cause was chest pain/dyspnea (codes: R06.0, R07.0, R07.1, R07.2, R07.3, or R07.4) and the individual had no record of a cardiac condition (all ICD cardiac related codes I.X). Exacerbations in general practitioner records (available for approximately 45% of the UK Biobank population) were identified using read v2 and read v3 codes for exacerbations (codes: H333, H3301, H3311, H33z0, H333z1, XE0YW, Xa1hD, Xafdy, Xafdz, Xafdj, XM0s2, 663d, or 8H2P, based on Shah et al.[28]). In the primary care prescription data, oral corticosteroids (OCS) prescriptions were analyzed for evidence of OCS bursts.[29, 30] OCS prescriptions were considered as exacerbations if the total dose prescribed was 200-600 mg. Prescriptions less than two weeks apart, less than 2 weeks after an annual asthma review (to avoid counting rescue packs) or less than 2 weeks after a general practitioner recorded exacerbation were excluded. Individuals not meeting any of these definitions were considered controls (non-exacerbators). To account for the potential that prescriptions may have suffered classification bias due to a fraction prescribed OCS bursts for other conditions, a sensitivity analysis was conducted restricting the definition of exacerbations to those identified using hospitalization data or primary care general practitioner diagnostic records. In GERA, exacerbations in the GERA cohort were defined as either an emergency department visit or hospitalization due to asthma (ICD10 codes: J45.XX, J46.XX), or an OCS burst prescription defined as a short course of oral steroids 3-21 days.

### Genotyping

Two arrays were used for genotyping: Affymetrix Axiom® UK BiLEVE array and the Affymetrix Axiom® UK Biobank array.[31] The Haplotype Reference Consortium panel was used for imputation. In total, 9,805,379 variants met our quality control criteria: minor allele frequency ≥ 0.05 and imputation score ≥ 0.3.

### Genome-wide analysis

A logistic regression model was used to determine genome-wide associations between genetic variants and asthma exacerbator status, assuming an additive genetic model. Imputed genotype dose (effect allele as a continuous variable ranging from 0-2, reflecting uncertainty in genotype imputation) were fitted using plink version 2.[32]. Age, sex, smoking status, the genotyping array, and the first 10 principal components (to adjust for population stratification) were included as covariates. To investigate effect modification by eosinophilia on the association between genetic variants and exacerbator status, we conducted a secondary analysis using eosinophilia (defined on a categorical basis using a cut-off of 300 cells/μl, collected during any of the three available visits to the UK Biobank assessment center) * variant as an interaction term. The Ensembl Variant Effect Predictor (VEP) was used to annotate the associated SNPs and analyze potential consequences.[33, 34] Fine-mapping was conducted using susieR package version 0.11.42.[35] All biallelic variants with MAF>0.0005 within a 1 Mb window surrounding the sentinel SNP were included. Summary statistics from logistic regression using Plink were passed to the susie_rss function with a variant correlation matrix generated from hard-calls using the cor function in R. Region plots were generated using LocusZoom.[36]

### Power analysis

Using GAS power calculator, detecting variants with a minimum relative risk of 1.3, with 80% power at a 5x10^-8^ significance level was estimated to require a sample size of at least 1,130 cases and 3,000 controls for risk alleles with a frequency of at least 10% (assuming frequency of exacerbators 40% among asthmatics).

## Results

### Stage 1

In total, 70,918 individuals were selected from the UK Biobank as asthma cases (based on self-reported asthma, hospitalization with an asthma code or a primary care record of asthma), of which 49,494 individuals met our inclusion criteria for this analysis. The baseline characteristics for all included individuals are shown in Table 1. Of included individuals, 11,604 (23.4%) met our criteria for having at least one exacerbation during follow-up (under any of the definitions used) and the remaining individuals (n=37,890) were considered as non-exacerbating controls. Exacerbators were slightly older and included a lower proportion of ever smokers.

**Table 1:** Baseline characteristics of cases and controls

| Characteristic | Exacerbators (n=11,604) | Non-exacerbators (n=37,890) |
| --- | --- | --- |
| Age | 57 ± 8 years | 56 ± 8 years |
| Female sex | 54.5% (n=7,096) | 54.2% (n=20,542) |
| Ever smoking | 52.2% (n=6,053) | 56.0% (n=21,231) |
| Eosinophilic (≥0.3 cells/μl) | 26.7% (n=3,117) | 23.5% (n=8,911) |
| Childhood onset asthma (<18 years at age of diagnosis) | 23.1% (n=2,680) | 29.3% (n=11,102) |
| Primary asthma hospitalization* | 14.4% (n=1,670) |  |
| Secondary hospitalization* | 6.2% (n=717) |  |
| Chest pain/Dyspnea*<sup>a</sup> | 13.0% (n=1,514) |  |
| Primary care (read codes)* | 9.8% (n=1,136) |  |
| Primary care exacerbations (prescriptions)* | 75.7% (n=8,788) |  |

\*=Some exacerbators met multiple definitions of exacerbations.
<sup>a</sup>= Secondary asthma hospitalizations with chest pain/dyspnea as primary hospitalization cause

**Table 2:** SNPs associated with asthma exacerbations within patients with asthma in both the UK Biobank and GERA

| Rsid | Allele | Chr:position | IS | Consequence | Nearest gene | MAF | OR | P value (stage 1) | OR (stage 2) | P value (stage 2) | OR (sensitivity analysis) | P value (sensitivity analysis) |
| --- | --- | --- | --- | --- | --- | --- | --- | --- | --- | --- | --- | --- |
| rs149721630 | C | 7: 48542408 | 0.94 | Intronic variant | *ABCA13* | 0.01 | 0.70 | 7.53 x10<sup>-8</sup> | 0.64 | 0.049 | 0.69 | 0.002 |
| rs34643691 | T | 15:88374252 | 0.94 | Intergenic variant | *NTRK3* | 0.01 | 1.46 | 8.67x 10<sup>-8</sup> | 1.74 | 0.017 | 1.46 | 0.003 |

OR= Odds Ratio, primary analysis= exacerbations defined using all sources of data, sensitivity analysis = exacerbations defined using hospitalization and primary care records data, MAF = Minor Allele Frequency, IS= Imputation info score

### Association signals for asthma exacerbations (stage 1)

No SNPs were associated with risk of asthma exacerbations at a genome-wide significance threshold (P<5x10^-8^), but 33 SNPs representing 12 independent loci were significant at a suggestive P value threshold (P<5x10^-6^). (Figure 1) A quantile-quantile plot is shown in Supplementary Figure 1. There were two suggestive signals (at P<5x10^-7^) with a minor allele frequency >1% in our population (Supplementary Table S3). These associations were followed up in a sensitivity analysis where exacerbations were defined as either hospitalizations or GP recorded exacerbations only (therefore excluding prescription data). The two SNPs retained a nominally significant association and direction of effect with exacerbations in the sensitivity analysis. Fine-mapping results showed both signals to be the most likely causal variant in their region. (Supplementary Figure 2)

**Figure 1:**
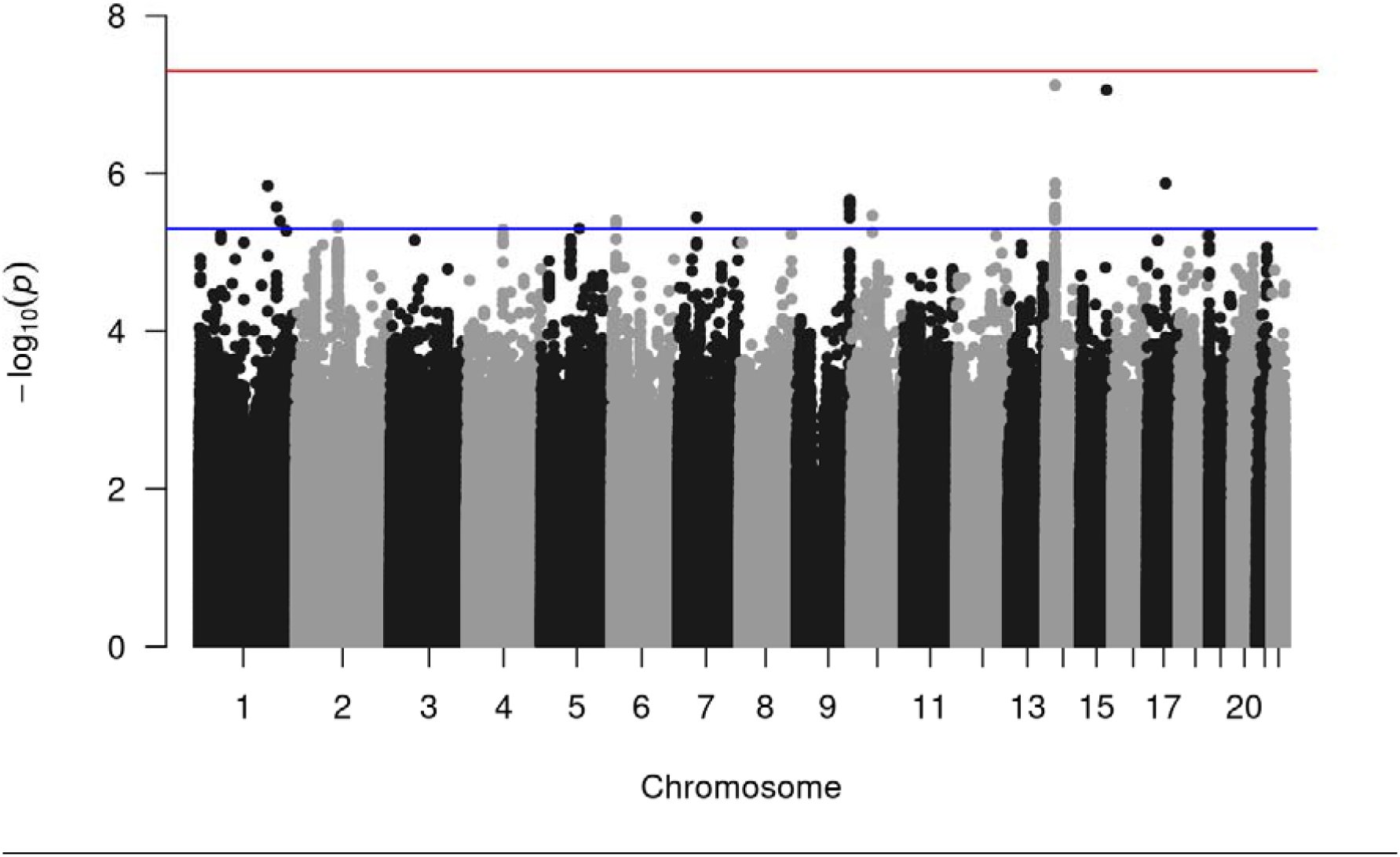
Manhattan plot of association results for asthma exacerbations in the UK Biobank. The x-axis shows genomic location by chromosome, the y-axis shows the –log10 P-value. The (lower) blue line indicates p= 5x10 ^-6^, and the (upper) red line corresponds to p= 5x10^-8^ (commonly known as genome-wide significance level).

### Association signals in stage 2 (GERA)

All signals showing a suggestive association in stage 1 (UK Biobank) were tested for association with exacerbations in an independent stage 2 study (GERA). Two variants (rs34643691 and rs149721630) showed consistent direction of effect and nominal significance (P<0.05) for association. (Supplementary file1).

### Regional associations

Figure 3 represents regional association plots for regions (±500 kb) centered at the two replicated SNPs. SNP rs34643691 is in an intergenic region close to Neurotrophic Receptor Tyrosine Kinase 3(NTRK3) and did not show strong linkage disequilibrium (LD) with other SNPs in the region. SNP rs149721630 is an intronic variant within ATP Binding Cassette Subfamily A Member 13 (ABCA13), and is in moderate-high LD with three other nearby SNPs.

**Figure 3:**
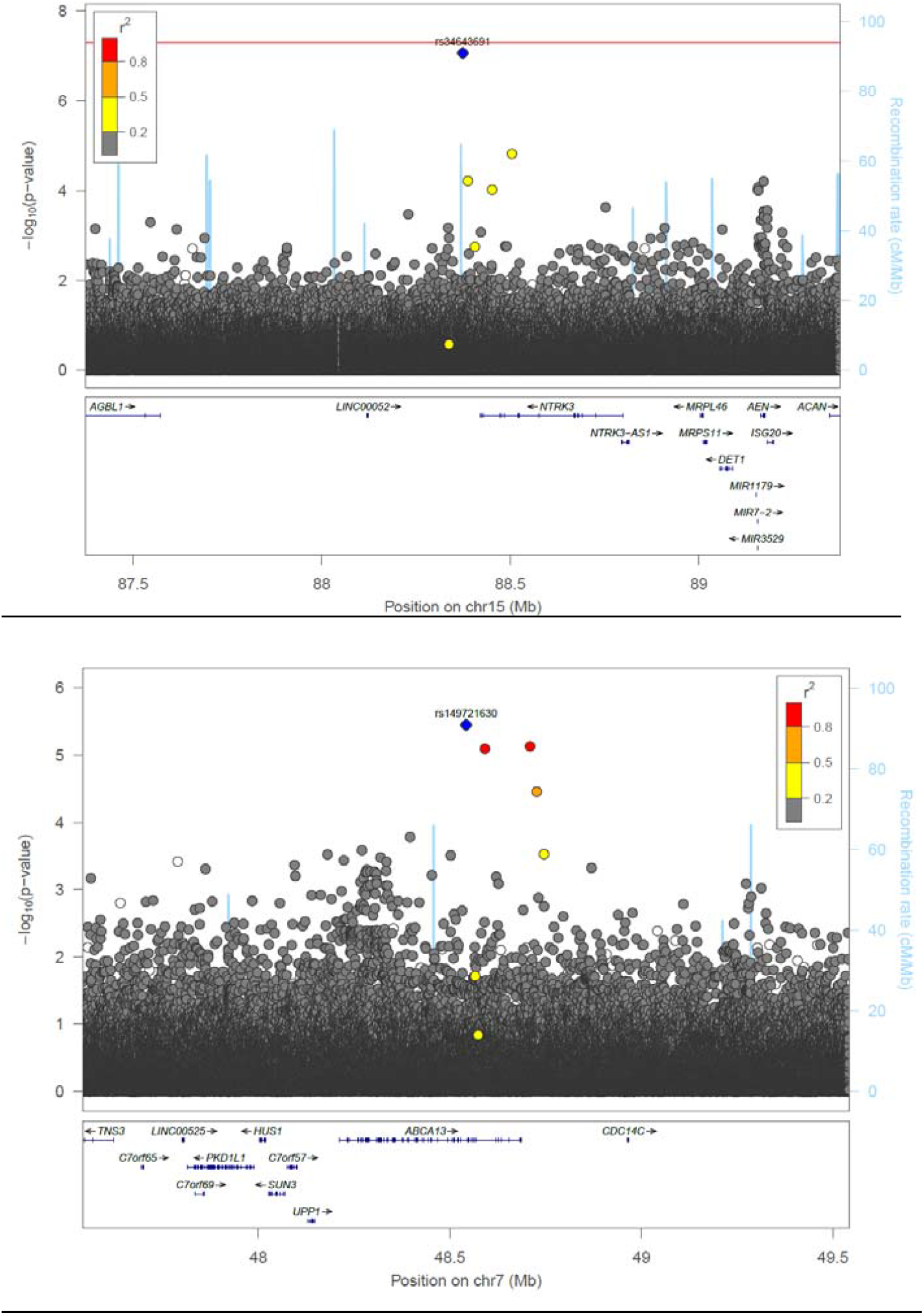
Region plots of replicated signals (A) rs34643691 and (B) rs149721630 results in stage 1. Plot is produced in LocusZoom. Blue diamond = sentinel SNP. Red= r^2^ greater than or equal to 0.8. Orange = r^2^ between 0.5 and 0.8. Yellow=r^2^ between 0.2 and 0.5. Grey= r^2^ less than 0.2. Genes are shown in the bottom panel in blue, with thicker sections representing exons.

### SNP by eosinophilia effect on exacerbations

Our interaction analysis showed no genome-wide significant associations and 86 signals that were associated with a gene by eosinophilia effect on exacerbations at P<5x10^-6^. The top hit comprised a locus on chromosome 18 overlapping two genes (*CXADRP3* and *POTEC*), as shown in the regional association plot (Figure 4). The minor allele of the sentinel variant increased exacerbation risk in eosinophilic patients (OR=1.17, P=0.0001) and decreased exacerbation risk in non-eosinophilic patients (OR=0.90, P=4.83x10^-5^).

**Figure 4:**
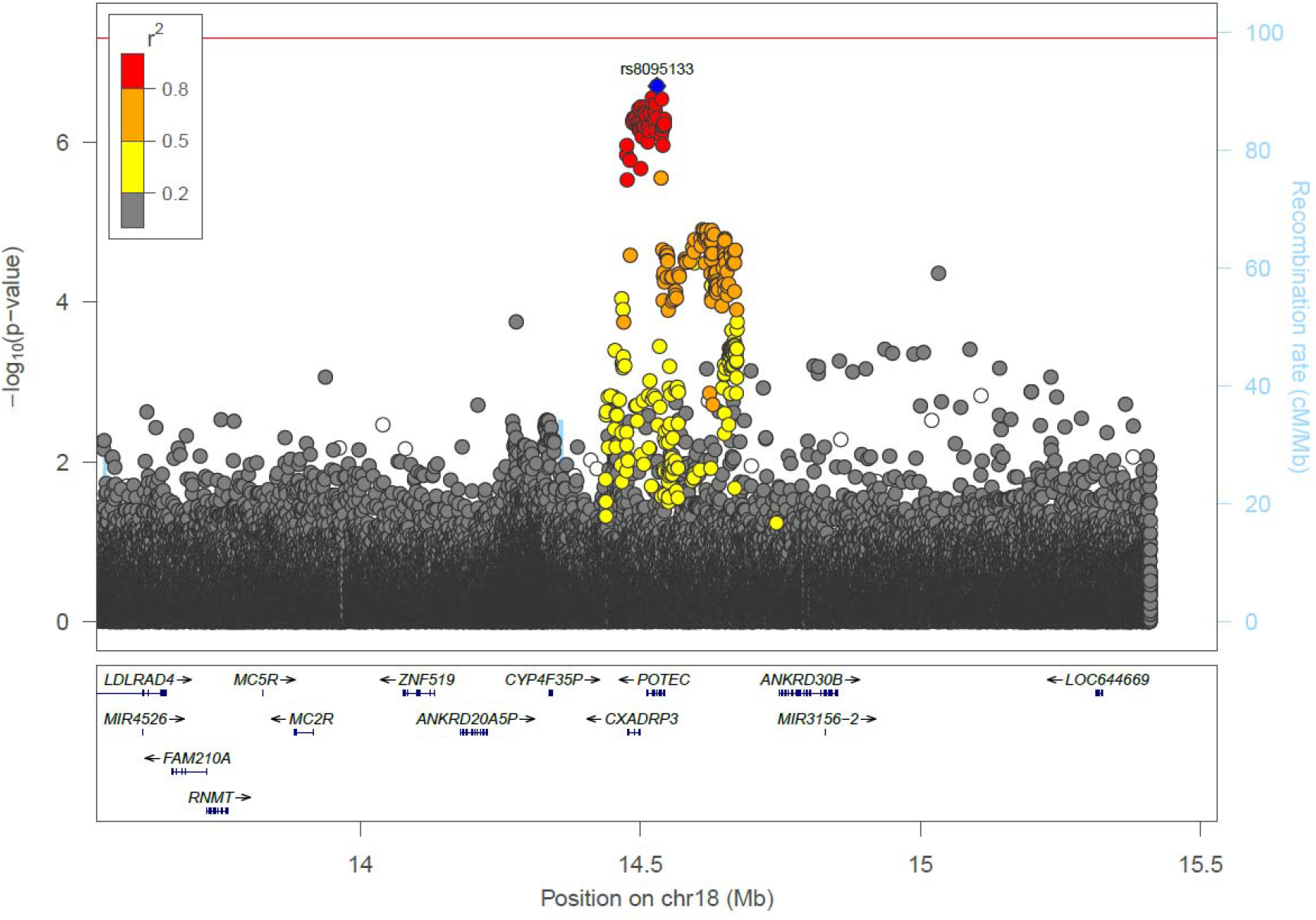
Genetic region of the chromosome 18 locus showing a significant interaction effect on exacerbation risk. The variants in strong LD are mainly associated with two genes, *CXADRP3* and *POTEC*. The top SNP (lowest p-value) in the regional analysis is rs8095133(POTEC). Blue diamond = sentinel SNP. Red= r^2^ greater than or equal to 0.8. Orange = r^2^ between 0.5 and 0.8. Yellow=r^2^ between 0.2 and 0.5. Grey= r^2^ less than 0.2. Genes are shown in the bottom panel in blue, with thicker sections representing exons.

### Downstream analyses

Using Haploreg and Ensembl Variant Effect Predictor, we annotated the top hits and identified potential functional consequences of SNPs in both steps of the analyses. [34] GWAS catalog analyses of the replicated associations, as well as the top hit in the interaction analysis did not reveal any previous associations with asthma. Additionally, the three SNPs did not show an eQTL effect on gene expression levels in the Genotype-Tissue Expression (GTEx) database.[37]

### Post-hoc power analysis

Aiming to confirm that we had enough power to detect significant associations, we conducted a post-hoc power analysis. The number of cases and controls included in our study (11604 and 37890, respectively) allows for the detection of variants with a minimum relative risk of 1.2 and an allele frequency of at least 0.05, with >80% power at a 5x10^-8^ significance level.

## Discussion

In this large, genome-wide association study, two novel loci were reproducibly associated with exacerbation risk in individuals with asthma. Exacerbations were identified using three different sources: hospitalization data, general practitioner records and OCS bursts prescriptions. In an interaction analysis aiming to explore effect modification of eosinophilia on the association between genetic variants and risk of asthma exacerbations, one locus showed opposite directions of effect on exacerbation risk in eosinophilic vs non-eosinophilic patients.

Two SNPs affected exacerbation status in our primary analysis: rs149721630 and rs34643691. Rs149721630 is an intron variant annotated to *ABCA13* gene, a member of the ATP-binding cassette (ABC) family of transmembrane transporters.[38] ABC (ATP-binding cassette) is a family of conserved transporters involved in transporting different substrates and consisting of seven sub-families.[38, 39] Members of the family have been previously associated with several lung conditions.[39, 40] *ABCA13* is expressed in the lungs and its expression levels in epithelial cells have been shown to be affected by asthma and smoking.[38] Additionally, decreased expression levels were associated with a trend towards increased asthma severity.[38] Moreover, differential methylation in the region was also associated with rhinovirus-induced wheezing and asthma.[41] Finally, *ABCA13* is associated with psychiatric disorders, including depression, a condition which shares potential links (including genetic) with asthma.[42, 43] This variant may indicate a potential link between this genetic region and asthma severity through an effect on impaired lipid transport or inflammation.[43, 44] However, this finding should be interpreted cautiously as it did not reach genome-wide significance in the discovery cohort. Conversely, rs34643691 lies in an intergenic region, and has not been previously associated to either regulatory elements or phenotypes and has a low minor allele frequency (1.03%). The closest gene is *NTRK3*, previously shown to be involved in neurotrophin signaling pathway, which may impact phosphatidyl inositol signaling via phosphatidylinositide 3-kinase (PI3K), affecting zileuton (a leukotriene receptor antagonist) related changes in Forced Expiratory Volume (FEV_1_).[45]

To investigate the hypothesis that there is effect modification by inflammatory subtype, we conducted an analysis using an interaction term (SNP x eosinophilia). Our interaction analysis identified an important signal on chromosome 18, associated with *CXDARP3* and *POTEC* genes. *CXDARP3* is a pseudogene for the CXDAR (Coxsackie Virus And Adenovirus Receptor) gene, and both are expressed in esophageal mucosal tissues.[46] This indicates a potential link between exacerbations in type 2 asthma and viral respiratory infections. Viral respiratory infections are well-known to be one of the most important triggers of asthma exacerbations, and adenoviruses can trigger wheezing and predispose to allergy.[47, 48] Importantly, viral infections and their interaction with allergens not only increase the risk of exacerbations, but also trigger an increased Th2 response.[49] Conversely, *POTEC* belongs to a multi-gene family encoding Cancer-Testis Antigens (CTAs), and is highly expressed in both ovaries and testes.[46, 50] While *POTEC* genes could constitute an important therapeutic target in cancer[50], it is possible that *POTEC* signals are significant in our analysis due to their high LD with *CXADRP3* variants. Alternatively, these variants could be playing regulatory roles within a complex network (e.g.: gene-gene interactions). Importantly, the SNPs identified in the initial analysis were not significant in the interaction analysis, suggesting that the effects of these genetic risk variants are not modified by background eosinophilia.

Our study has several unique strengths: first, we conducted and replicated our GWAS of asthma exacerbations within adults, whereas most previous studies have been conducted in children. Second, we focused on the potential differences between genetic variants affecting the general risk of asthma exacerbations and variants affecting exacerbations modified by type 2 asthma (defined as patients with a blood eosinophil level ≥300 cells/μl). This is a unique approach which allows unraveling variants affecting exacerbations in individuals with specific active inflammatory pathways. Finally, we used a broad definition of exacerbations, by including data from various sources in both analysis stages. We included severe exacerbations (hospitalizations) using several definitions, aiming for a wider coverage of hospitalizations associated with asthma. Hospitalizations (in individuals with asthma) recorded as primarily due to influenza, respiratory failure, stridor, wheezing or respiratory failure were considered asthma exacerbations. Hospitalizations associated with chest pain or dyspnea in non-cardiac individuals with asthma were also considered exacerbations. Additionally, we added exacerbations in the community using prescription data and general practitioner records. This broad definition of asthma exacerbations aimed to provide a more accurate picture of asthma exacerbations by including all asthma exacerbators. Using this approach, 23.5% of our asthma population had at least one exacerbation, a figure close to previous estimates indicating the potential usefulness of the broader definition.[5] As most exacerbators were identified based on OCS prescriptions, we carried out sensitivity analyses to show that these variants had consistent magnitude and direction of effect in those identified from diagnostic ICD-10 and primary care read codes alone.

However, our study also had several limitations. First of all, there were no genome-wide significant associations in our primary analysis of exacerbation risk or in our interaction analysis within adult asthmatics. This could be in part due to power, especially as most of our top hits had a relatively low minor allele frequency.[51] Second, our analysis was restricted to individuals of European ancestry and therefore our results may not be generalizable to other populations. Third, our definition of eosinophilic patients was only based on blood eosinophil levels taken during any of three visits to the UK biobank assessment center. Airway eosinophilia could have provided a more accurate definition of eosinophilic patients, but this is less feasible in population-wide studies.[52] Moreover, eosinophilia in participants with less visits to the centers could have been underestimated. Fourth, we only assumed an additive genetic model, while some SNPs could be affecting asthma under different genetic models. Fifth, we did not include follow-up time in our model. In the UK Biobank, hospitalization data was available from 1995 up to 2020, while primary care data (for half the cohort) was available from 1995 until 2016, which could have caused differences in follow-up and therefore the ability to record an exacerbation. Sixth, we did not replicate the interaction analysis assessing the effects of eosinophilia on the association between SNPs and asthma exacerbations, as blood eosinophil levels were not available in the GERA cohort. Finally, there could be three main criticisms to our definition of asthma exacerbations: first, we defined all chest pain/dyspnea in asthmatic non-cardiac patients admitted to a hospital as exacerbations. We aimed to include important signs of exacerbations which would be otherwise missed due to the strict coding system, as these symptoms may be confusing to clinicians on a patient’s initial presentation, and are common in acute asthma.[53] Second, UK Biobank secondary care data does not include emergency department admissions unless those patients are transferred as in-patients to another department. We will therefore have missed any such exacerbations. Finally, we used OCS burst prescriptions to identify asthma exacerbations in the community setting, which could be difficult to fully ascertain as patients may have had additional comorbidities for which they were prescribed OCS at the same dose (also as the available data on the intended duration of the specific prescription was missing). Alternatively, clinicians might have used lower doses intended for exacerbations than the dose threshold we used. Both broad definitions could have misclassified some patients as exacerbators or controls. However, the moderate percentage of exacerbators within our population, as well as the results of sensitivity analyses suggest that any potential misclassification had limited effects on our final results.

Understanding the complex factors affecting the severity of asthma is essential to advance the understanding and treatment of asthma. Our study adds to the existing evidence of genetic involvement in the severity of asthma and the presenting clinical phenotype. Further work to translate findings and understand exactly how these variations are contributing towards different inflammatory pathways is required. Additional layers of data (including epigenetic or proteomics data) may be needed to unravel the biologically complex network of interactions, aiming to identify disease markers and/or drug targets.

In conclusion, our GWAS in the UK biobank identified two variants that are associated with the risk of exacerbations, highlighting a potentially important effect for *ABCA13*. Additionally, we identified a locus associated with *CXADRP3* and *POTEC* genes affecting exacerbation risk only in eosinophilic patients. These findings could shed light on important pathways involved in asthma severity.

## Data Availability

Summary statistics will be made available upon reasonable request to the authors

---

**Supplementary Figure 1:**
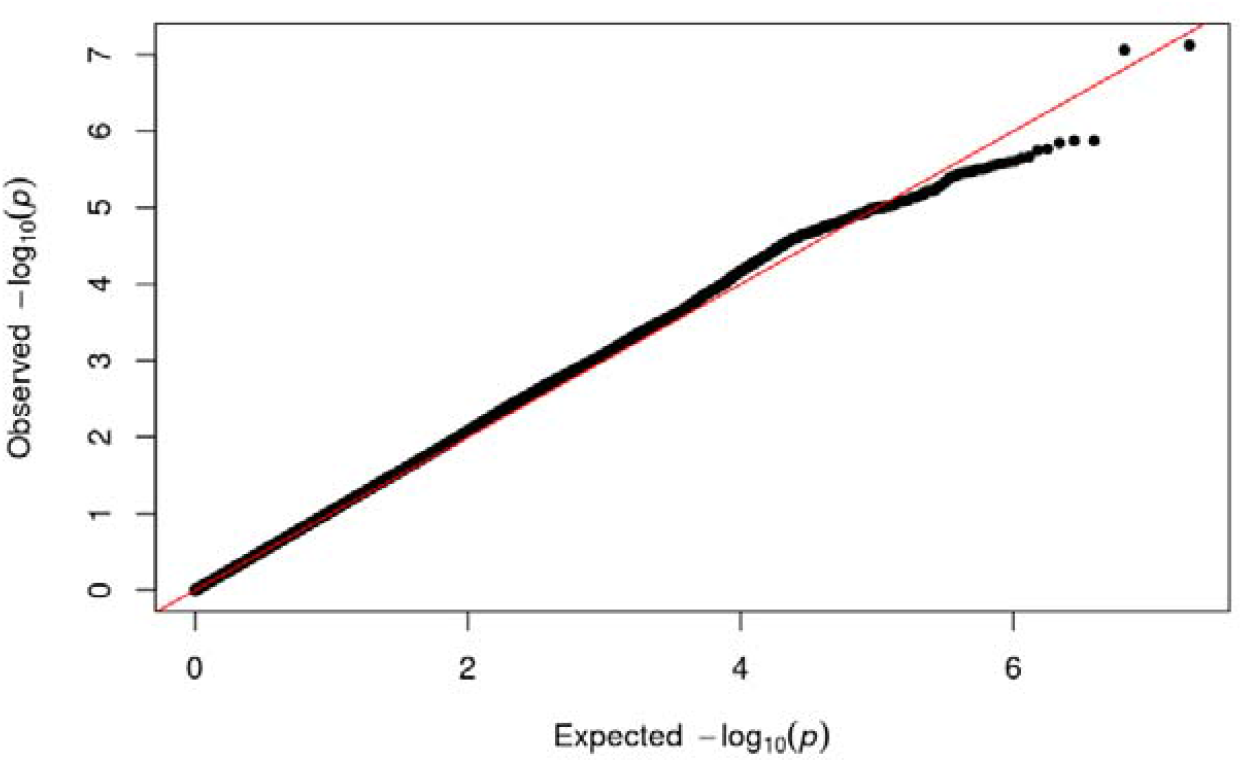
Quantile-quantile plot of genome-wide associations of asthma exacerbations within asthmatics (λ=1.043)

**Supplementary Figure 2:**
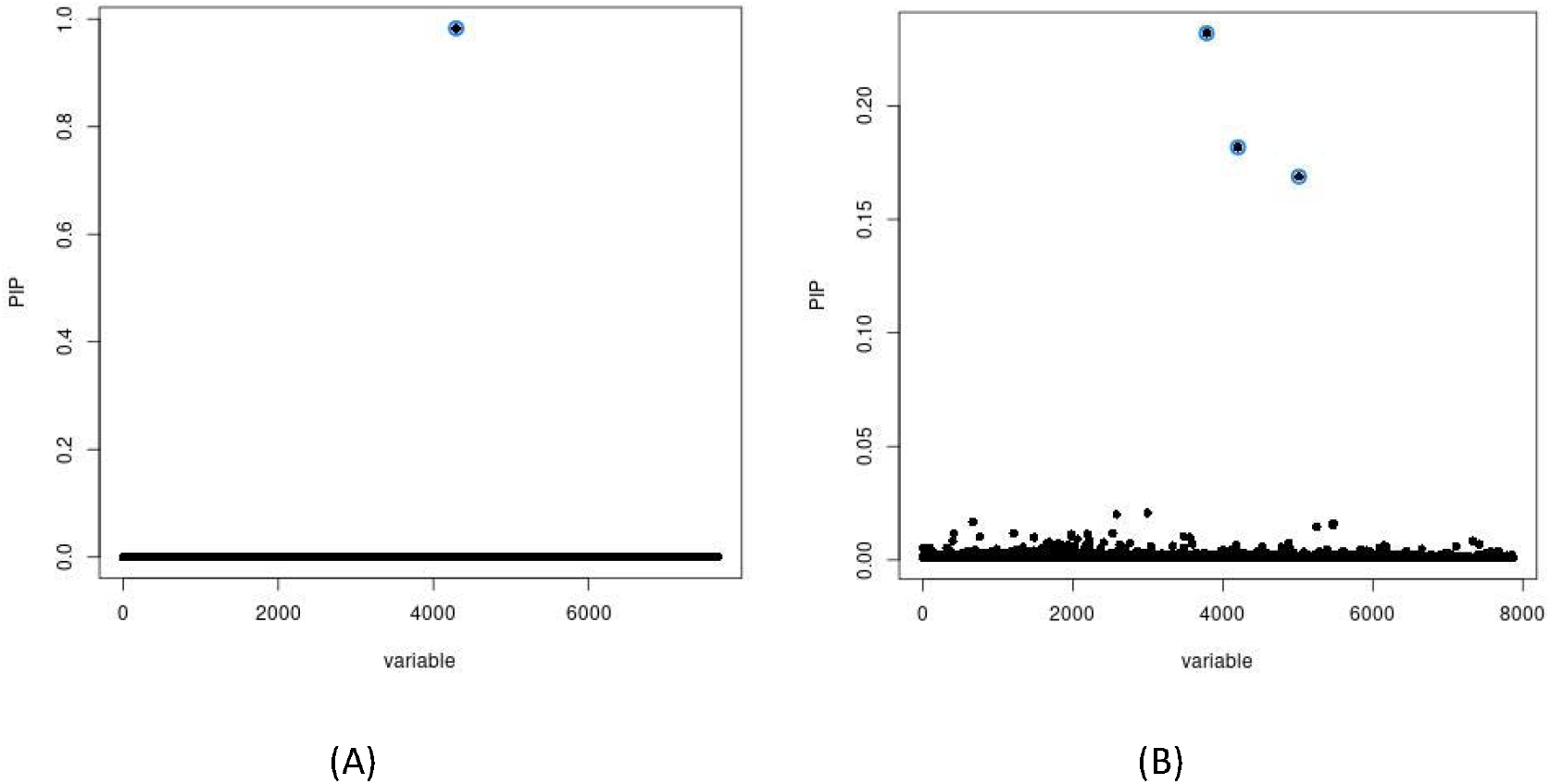
Credile sets for (A) rs34643691 and (B) rs149721630

**Supplementary Table S3.**
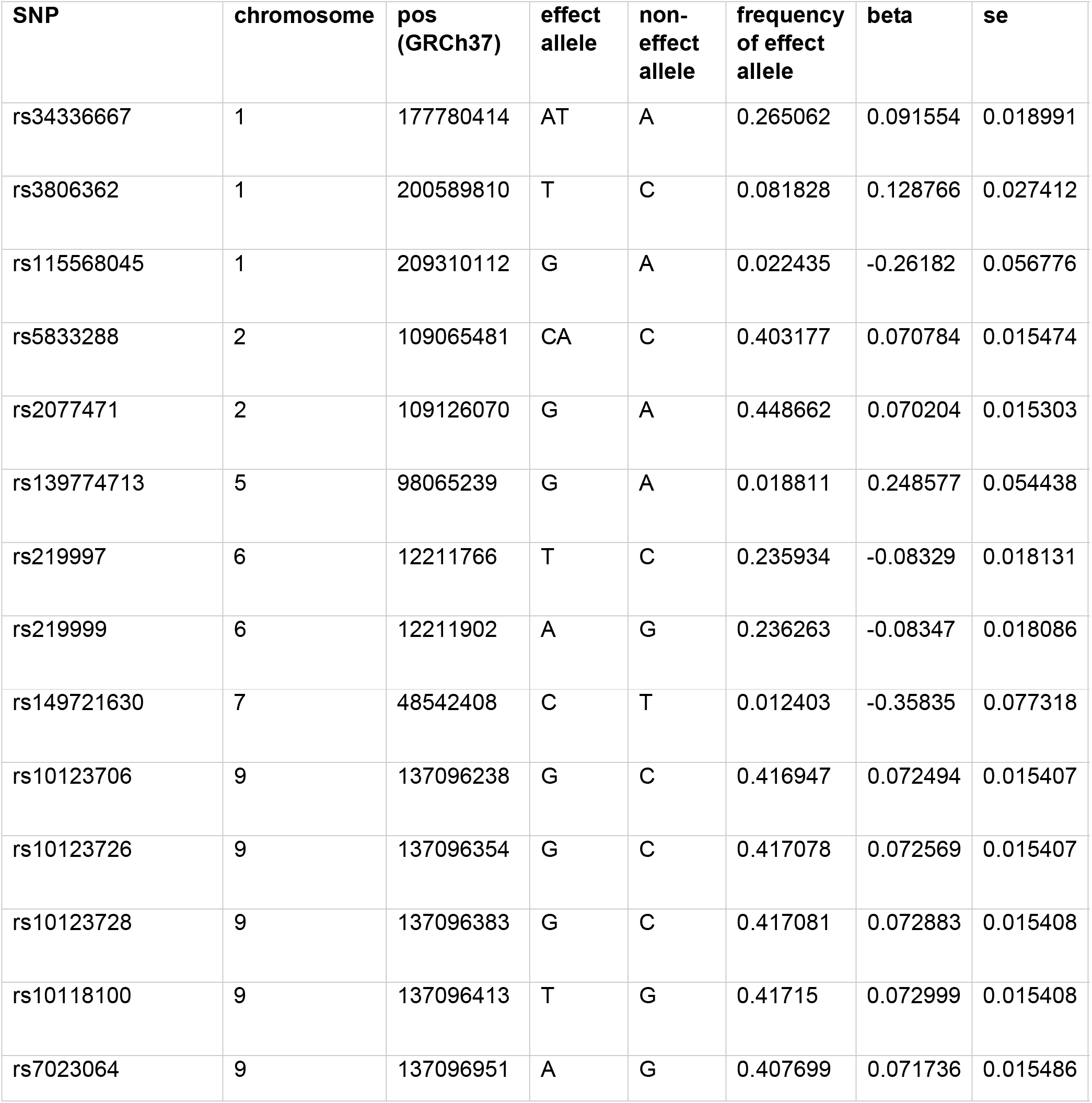
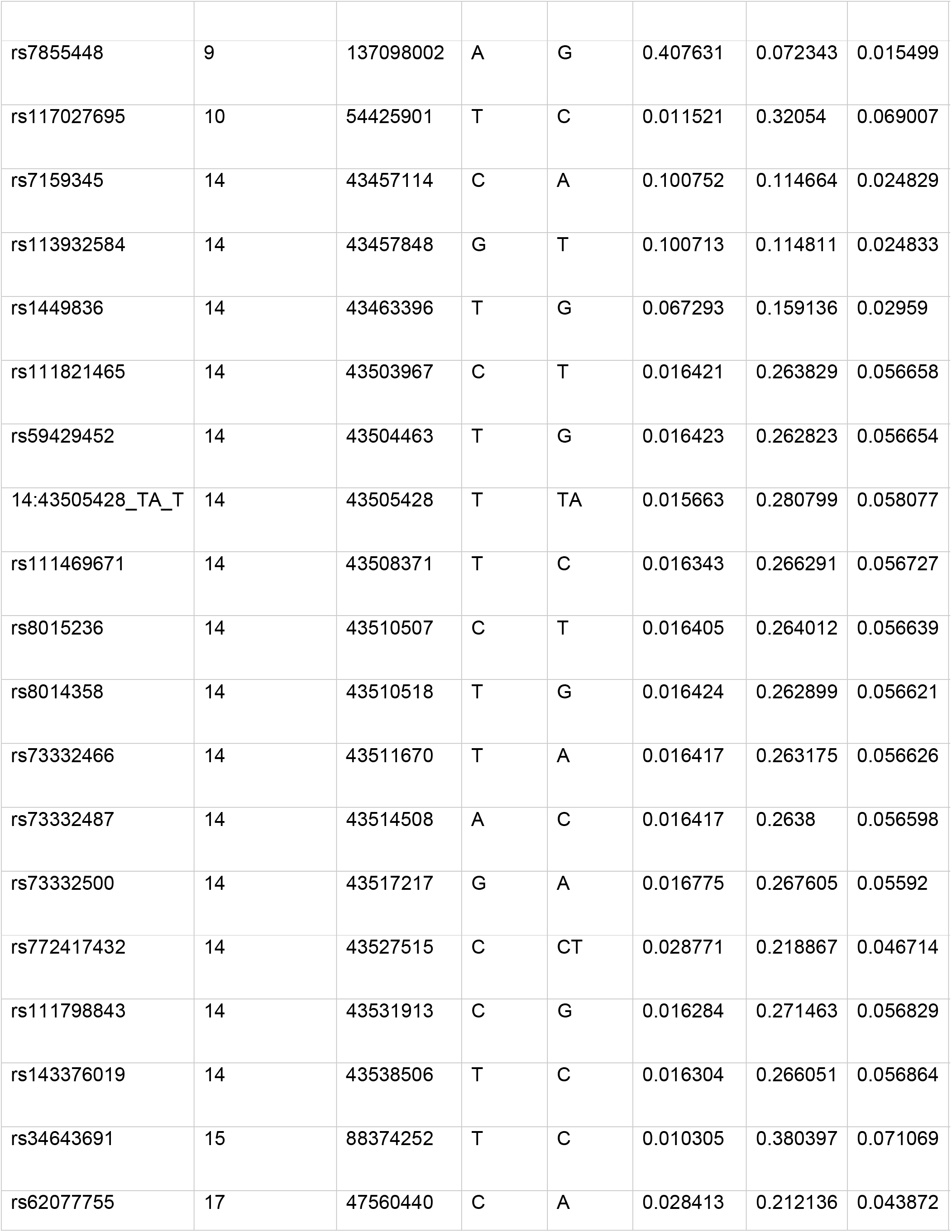
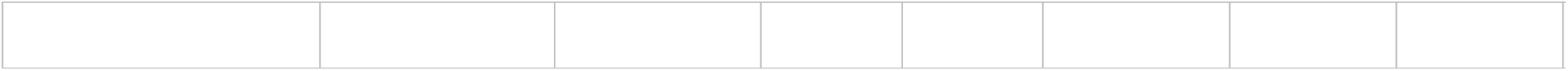

| SNP | chromosome | pos<br>(GRCh37) | effect<br>allele | non-<br>effect<br>allele | frequency<br>of effect<br>allele | beta | se |
| --- | --- | --- | --- | --- | --- | --- | --- |
| rs34336667 | 1 | 177780414 | AT | A | 0.265062 | 0.091554 | 0.018991 |
| rs3806362 | 1 | 200589810 | T | C | 0.081828 | 0.128766 | 0.027412 |
| rs115568045 | 1 | 209310112 | G | A | 0.022435 | -0.26182 | 0.056776 |
| rs5833288 | 2 | 109065481 | CA | C | 0.403177 | 0.070784 | 0.015474 |
| rs2077471 | 2 | 109126070 | G | A | 0.448662 | 0.070204 | 0.015303 |
| rs139774713 | 5 | 98065239 | G | A | 0.018811 | 0.248577 | 0.054438 |
| rs219997 | 6 | 12211766 | T | C | 0.235934 | -0.08329 | 0.018131 |
| rs219999 | 6 | 12211902 | A | G | 0.236263 | -0.08347 | 0.018086 |
| rs149721630 | 7 | 48542408 | C | T | 0.012403 | -0.35835 | 0.077318 |
| rs10123706 | 9 | 137096238 | G | C | 0.416947 | 0.072494 | 0.015407 |
| rs10123726 | 9 | 137096354 | G | C | 0.417078 | 0.072569 | 0.015407 |
| rs10123728 | 9 | 137096383 | G | C | 0.417081 | 0.072883 | 0.015408 |
| rs10118100 | 9 | 137096413 | T | G | 0.41715 | 0.072999 | 0.015408 |
| rs7023064 | 9 | 137096951 | A | G | 0.407699 | 0.071736 | 0.015486 |
| rs7855448 | 9 | 137098002 | A | G | 0.407631 | 0.072343 | 0.015499 |
| rs117027695 | 10 | 54425901 | T | C | 0.011521 | 0.32054 | 0.069007 |
| rs7159345 | 14 | 43457114 | C | A | 0.100752 | 0.114664 | 0.024829 |
| rs113932584 | 14 | 43457848 | G | T | 0.100713 | 0.114811 | 0.024833 |
| rs1449836 | 14 | 43463396 | T | G | 0.067293 | 0.159136 | 0.02959 |
| rs111821465 | 14 | 43503967 | C | T | 0.016421 | 0.263829 | 0.056658 |
| rs59429452 | 14 | 43504463 | T | G | 0.016423 | 0.262823 | 0.056654 |
| 14:43505428_TA_T | 14 | 43505428 | T | TA | 0.015663 | 0.280799 | 0.058077 |
| rs111469671 | 14 | 43508371 | T | C | 0.016343 | 0.266291 | 0.056727 |
| rs8015236 | 14 | 43510507 | C | T | 0.016405 | 0.264012 | 0.056639 |
| rs8014358 | 14 | 43510518 | T | G | 0.016424 | 0.262899 | 0.056621 |
| rs73332466 | 14 | 43511670 | T | A | 0.016417 | 0.263175 | 0.056626 |
| rs73332487 | 14 | 43514508 | A | C | 0.016417 | 0.2638 | 0.056598 |
| rs73332500 | 14 | 43517217 | G | A | 0.016775 | 0.267605 | 0.05592 |
| rs772417432 | 14 | 43527515 | C | CT | 0.028771 | 0.218867 | 0.046714 |
| rs111798843 | 14 | 43531913 | C | G | 0.016284 | 0.271463 | 0.056829 |
| rs143376019 | 14 | 43538506 | T | C | 0.016304 | 0.266051 | 0.056864 |
| rs34643691 | 15 | 88374252 | T | C | 0.010305 | 0.380397 | 0.071069 |
| rs62077755 | 17 | 47560440 | C | A | 0.028413 | 0.212136 | 0.043872 |

